# Serine/threonine kinase activity associates with brain glucose metabolism changes in Alzheimer’s Disease

**DOI:** 10.1101/2022.10.31.22281751

**Authors:** Guilherme Povala, Marco Antônio De Bastiani, Bruna Bellaver, Pamela C. L. Ferreira, João Pedro Ferrari-Souza, Firoza Z. Lussier, Diogo O. Souza, Pedro Rosa-Neto, Bruno Zatt, Tharick A. Pascoal, Eduardo R. Zimmer, the Alzheimer’s Disease Neuroimaging Initiative

## Abstract

**Background:** Positron emission tomography (PET) imaging has greatly improved the diagnosis and monitoring of Alzheimer’s disease (AD). The recently developed neuroinformatic field is expanding analytical and computational strategies to study multimodal neuroscience data. One approach is integrating PET imaging and omics to provide new insights into AD pathophysiology.

**Methods:** Hippocampal and blood transcriptomic data of cognitively unimpaired (CU) and cognitively impaired (CI) individuals were obtained from Gene Expression Omnibus (GEO) databases and the Alzheimer’s Disease Neuroimaging Initiative (ADNI). We used the differentially expressed genes (DEGs) from these datasets to implement a modular dimension reduction approach based on Gene Ontology (GO) and reverse engineering of transcriptional networks centered on transcription factors (TF). GO clusters and regulatory units of TF were selected to undergo integration with [^18^F]Fluorodeoxyglucose ([^18^F]FDG)-PET images using voxel-wise linear regression models adjusted for age, gender, years of education, and *APOE* ε4 status.

**Results:** The GO semantic similarity resulted in 16 GO clusters enriched with overlapping DEGs in blood and the brain. Voxel-wise analysis revealed a strong association between the cluster related to the regulation of protein serine/threonine kinase activity and the [^18^F]FDG-PET signal in the brain. The master regulator analysis showed 61 regulatory units of TF significantly enriched with DEGs. The voxel-wise analysis of these regulons showed that zinc-finger-related regulatory units had the closest association with brain glucose metabolism.

**Conclusion:** We identified multiple biological processes and regulatory units of TF associated with [^18^F]FDG-PET metabolism in the brain of individuals across the aging and AD clinical spectrum. Furthermore, the prominent enrichment of protein serine/threonine kinase activity-related GO cluster and the zinc-finger-related regulatory units highlight the potential gene signatures associated with changes in glucose metabolism due to AD pathology.

## INTRODUCTION

Alzheimer’s disease (AD) is the most prevalent neurodegenerative disease affecting the elderly (1). In recent decades, a vital concept shift has led to AD being recognized as a clinical-biological entity. In this context, high-resolution magnetic resonance imaging (MRI) and positron emission tomography (PET) currently allow for the visualization of AD pathological changes in living individuals (2). Indeed, these technologies have highly contributed to an improved understanding of AD and boosted a new era of enormous imaging data collection in multiple centers worldwide and collaborative initiatives.

The multifaceted nature of AD requires the incorporation of complex paradigms to study its origins, mechanisms, development, and possible treatments. In this context, systems biology methods can be extremely useful in unraveling disease complexity (3,4). A systems view of biology describes a multidisciplinary and holistic approach to understanding biological phenomena, focusing on the interactions of many elements simultaneously (5,6). Among many possibilities, omics technologies, such as transcriptomics, are the methods of choice to quickly, broadly, and reliably evaluate systems properties and dynamics. Transcriptomics is especially attractive because RNAs represent an intermediate level between a static genomics context and the spatiotemporal dynamic of proteomics complexity (7).

There has been an ever-increasing call for data integration in recent years. The systems biology field culminated in current trends toward multi-omics approaches to generate models of hierarchical biological networks and changes in the system as it moves from a normal to a pathological condition (8–11). In neurosciences, these concepts and ideas resulted in the neuroinformatic subfield devoted to developing analytical and computational models for sharing, integrating, and analyzing multimodal neuroscience data. In this context, strategies to integrate neuroimaging data with transcriptomics started to emerge at the gene level (12–14). However, genes rarely dictate phenotypes alone, acting in concert to modulate phenotypic transitions.

In the present study, we sought to implement two modular, systems-based strategies to integrate the transcriptomics information of groups of genes with PET neuroimaging data in AD. In the first, clusters of Gene Ontology (GO) terms grouped by semantic similarity were associated with [^18^F]FDG-PET images. The second strategy used reverse engineering of transcriptional networks to integrate regulatory units of transcription factors with [^18^F]FDG-PET images. We hypothesize that modular approaches based on groups of genes will yield better associations with higher-order biological manifestations and highlight important AD metabolic and molecular features.

## METHODS

### Blood Expression Data Acquisition

We obtained blood microarray data of 743 individuals from the Alzheimer’s Disease Neuroimaging Initiative (ADNI) (http://adni.loni.usc.edu/). We included samples with RNA integrity (RIN) scores above seven in the study. After RIN and quality control filters, the resulting ADNI dataset retained 317 samples [218 cognitively impaired (CI) and 99 cognitively unimpaired (CU) individuals]. We obtained additional processed blood microarray datasets from Gene Expression Omnibus (GEO) repository (https://www.ncbi.nlm.nih.gov/geo/). GSE63063 (15) contains 433 CI and 223 CU samples from AddneuroMed Cohort, and GSE97760 (16) includes 8 CI and 7 CU controls (**Figure 1A**). Metadata from these datasets is shown in **Supplemental Table 1**.

**Figure 1.**
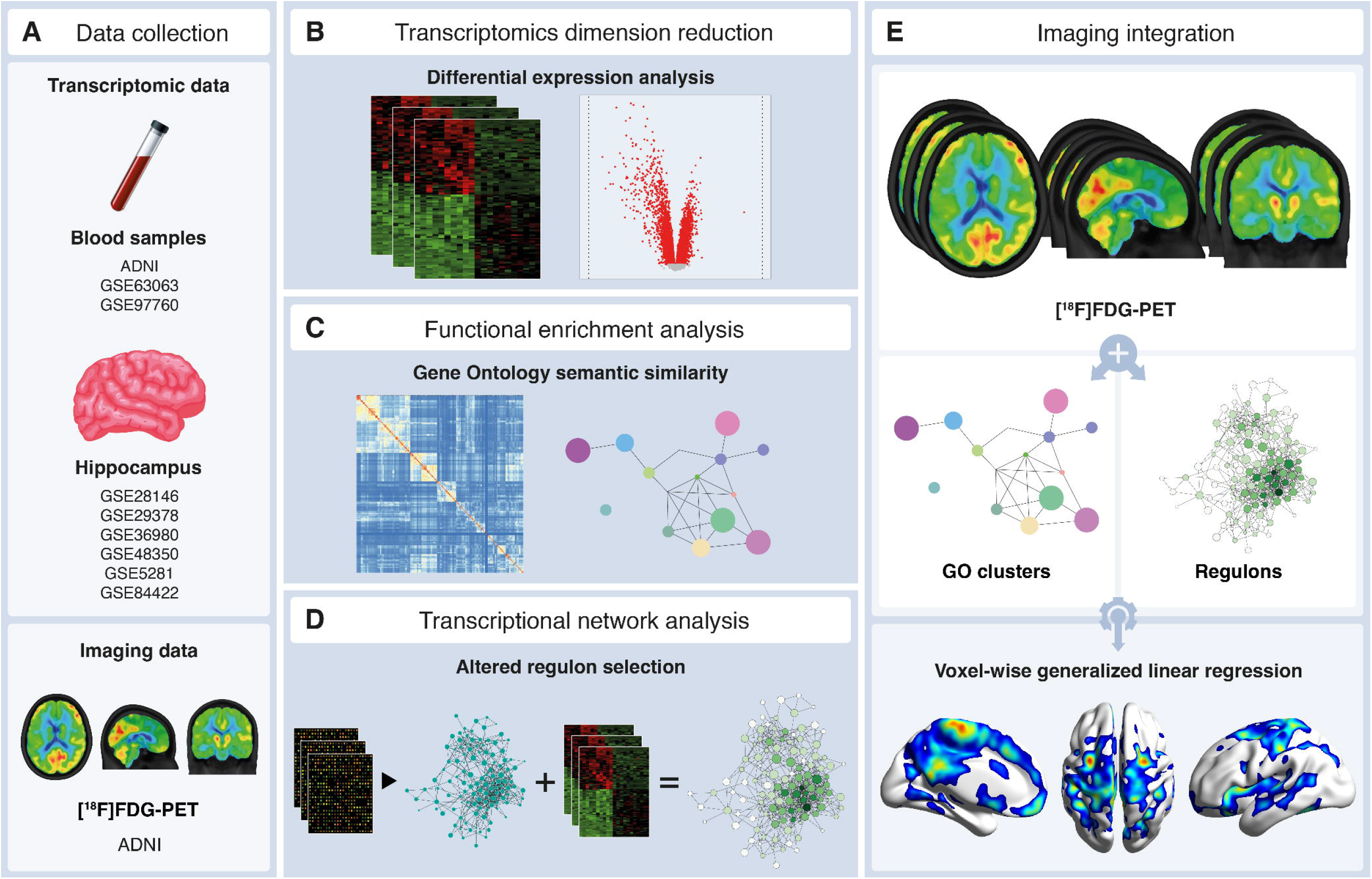
Omics-imaging integration pipeline. (A) Data collection of blood and hippocampus samples and imaging data from multiple datasets. (B) Transcriptomics dimension reduction using differential expression analysis. (C) Functional enrichment analysis using the Gene Ontology (GO) semantic similarity. (D) Transcriptional network analysis, responsible for selecting altered regulatory units. (E) Imaging integration using voxel-wise generalized linear regression between PET images and statistically altered GO clusters and regulatory units.

Healthy human blood samples from two large microarray datasets were obtained from the GEO repository [GSE48348 (17) and GSE99039 (18)], downloaded using the *GEOquery* package (v2.56.0) (19), and combined under common gene symbol annotations using the *virtualArray* package (v1.8.0) (20). The resulting dataset retained 967 healthy blood samples used for transcriptional network inference.

### Brain Expression Data Acquisition

Human AD hippocampal microarray data of six studies were obtained from GEO repository [GSE5281 (21), GSE28146 (22), GSE29378 (23), GSE36980 (24), GSE48350 (25) and GSE84422 (26)], downloaded using the *GEOquery* package (v2.56.0) (19) and combined under common gene symbol annotations using the *virtualArray* package (v1.8.0) (20) (**Figure 1A**). We processed raw CEL data using the *affy* package (v1.70.0) (27), and RMA was corrected using the *vsn* package (v3.60.0) (28). Afterward, we implemented a batch correction using the *sva* package (v3.40.0) (29). The combined dataset contained 90 CI and 116 CU individuals (**Supplemental Table 1**). Sample demographics can be found in **Supplemental Table 2**.

### Differential Expression Analyses

Differential expression analyses were computed on microarray data using the *limma* package (v3.48.3) (30) *lmFit* function to fit multiple linear models by generalized least squares. In addition, *eBayes* function was used to compute moderated t-statistics, moderated F-statistics, and log-odds of differential expression by empirical Bayes moderation of the standard errors towards a common value (**Figure 1B**).

We reasoned that brain expression would be more sensitive to alterations than blood expression in AD. Thus, genes with an unadjusted p-value < 0.05 in at least ⅔ of the tested datasets were considered differentially expressed genes (DEGs) for blood samples. While, for brain samples, genes with FDR adjusted p-value < 0.01 from the merged dataset were considered DEGs. We obtained the final list of genes used for further analyses from the intersection between blood and brain DEGs. Venn diagrams were constructed using the *VennDiagram* package (v1.6.20) (31). Details about differentially expressed genes from datasets are shown in **Supplemental Tables 3 and 4**.

### Functional Enrichment Analyses and Gene Ontology Semantic Similarity

DEGs intersecting blood between brain datasets were submitted to Gene Ontology (GO) enrichment analysis using the *clusterProfiler* package (v4.0.5) (32) *enrichGO* function. We clustered the GO biological process terms by semantic similarity using the *mgoSim* function from the *GOSemSim* (v2.18.1) package (33) (arguments measure = “Wang’’ and combine = NULL). Semantic comparisons of Gene Ontology (GO) annotations provide quantitative ways to compute similarities between genes and gene groups. We represented the resulting similarity matrices as GO networks using the *RedeR* (v1.40.0) package (34) for interactive visualization and manipulation of nested networks. We retained only similarity scores above the 80th percentile of score distribution for building the network. We manually named clusters of GO terms obtained from the GOSemSim algorithm for their biological interpretation (**Figure 1C**).

### Reverse Engineering of Transcriptional Network and Master Regulator Analysis

The transcriptional network (TN) centered on transcription factors (TF), and their predicted target genes were inferred using a large cohort of healthy blood samples and merged as described in **Blood Expression Data Acquisition**. Herein, we used “regulatory unit” or “regulon” to describe the groups of inferred genes and their associated TFs. *RTN* (v2.16.1) package was used to reconstruct and analyze TNs based on the mutual information (MI) using the Algorithm for the Reconstruction of Accurate Cellular Networks (ARACNe) method (35–37). In summary, the regulatory structure of the network is derived by mapping significant associations between known TFs and all potential targets. A curated list of genes obtained from (38) was used to annotate TF eligibility for TN inference input. To create a consensus bootstrap network, we eliminated the interactions below a minimum MI threshold by a permutation step and removed unstable interactions by bootstrap. Finally, the data processing inequality algorithm is applied with null tolerance to eliminate interactions likely to be mediated by a third TF. The reference blood TN was built using the package’s default number of 5000 permutations and 100 bootstraps (p-value < 0.001).

We conducted the master regulator analysis (MRA) described by Carro and colleagues (39) on the regulatory units with more than 100 targets using the RTN package. For each regulatory unit in the blood TN, the algorithm computes the statistical overrepresentation (calculated by Fisher’s exact test) of genes obtained from differential expression analyses (unadjusted p-value < 0.05). Master regulator candidates altered in at least ⅔ of the three case-control studies (ADNI, GSE63063, and GSE99039) were selected for gene set variation analysis and neuroimaging integration (**Figure 1D**).

### Gene Set Variation Analysis

The enrichment scores (ES) obtained from the gene set variation analysis (GSVA) method were used to collapse the activity of groups of genes into a single value for each sample. This gene set enrichment method estimates group gene activity variation over a sample population in an unsupervised manner. Briefly, genes contained in the clusters of GO terms obtained from GOSemSim or in the regulatory units obtained from master regulator analysis were submitted to the *gsva* function (kcdf=“Gaussian”) of the *GSVA* package in R (40). We used the resulting ES for further association with neuroimaging data and correlation analyses.

### Imaging pre-processing and integration

We acquired imaging data were ADNI (**Figure 1A**), and we downloaded [^18^F]FDG-PET images in the “*Co-reg, avg, std img and vox size, uniform resolution*” pre-processed format. We downloaded MPRAGE MRI images with “*gradwarp, B1 non-uniformity, and N3 image*” pre-processing correction steps. [^18^F]FDG-PET images were linearly registered to the T1-weighted image space. We linearly and nonlinearly registered T1-weighted images to the MNI template space. Subsequently, we performed an [^18^F]FDG-PET nonlinear registration using the linear and nonlinear transformations from the T1-weighted image to the MNI space and the [^18^F]FDG-PET to T1-weighted image registration. We performed these transformations using the *antsRegistrationSyNQuick* and *antsApplyTransforms* from advanced normalization tools (ANTs) (41). We spatially smoothed the [^18^F]FDG-PET images to achieve a final resolution of 8mm full width at half maximum using *mincblur* from MINC Tools (http://bic-mni.github.io/).

We generated the [^18^F]FDG-PET standardized uptake ratio (SUVr) maps using the cerebellum as a reference region. GO clusters and regulons obtained in the previous step were selected to undergo integration with [^18^F]FDG-PET images using voxel-wise linear regression models adjusted for age, gender, and years of education (RMINC package) (https://github.com/Mouse-Imaging-Centre/RMINC) (**Figure 1E**). The voxel-wise correlations resulted in t-statistical maps. Afterwards, only statistically significant correlated voxels (| t-value | > 2.6 [p-value of 0.01]) were retained.

## RESULTS

The dimension reduction strategy proposed in this work is represented by the Sankey flow diagram in **Figure 2**. Specifically, the union between blood and hippocampus transcriptomic data resulted in 16972 unique genes. When comparing CU and CI individuals from the 16126 blood genes, the differential expression analysis showed 2875 DEGs (unadjusted p-value < 0.05 present in at least ⅔ of datasets). From the 10845 hippocampus genes, we found 1979 DEGs (FDR adjusted p-value < 0.01 from the merged dataset) in comparing CU and CI individuals. Considering the DEGs from blood and hippocampus, 428 were consistently altered between tissues. FEA of Gene Ontology using these genes revealed 89 terms significantly enriched with altered elements (**Supplemental Table 5**). These results showed consistent modifications in functional terms associated with oxidative phosphorylation in mitochondria and cell cycle and response to stress.

**Figure 2.**
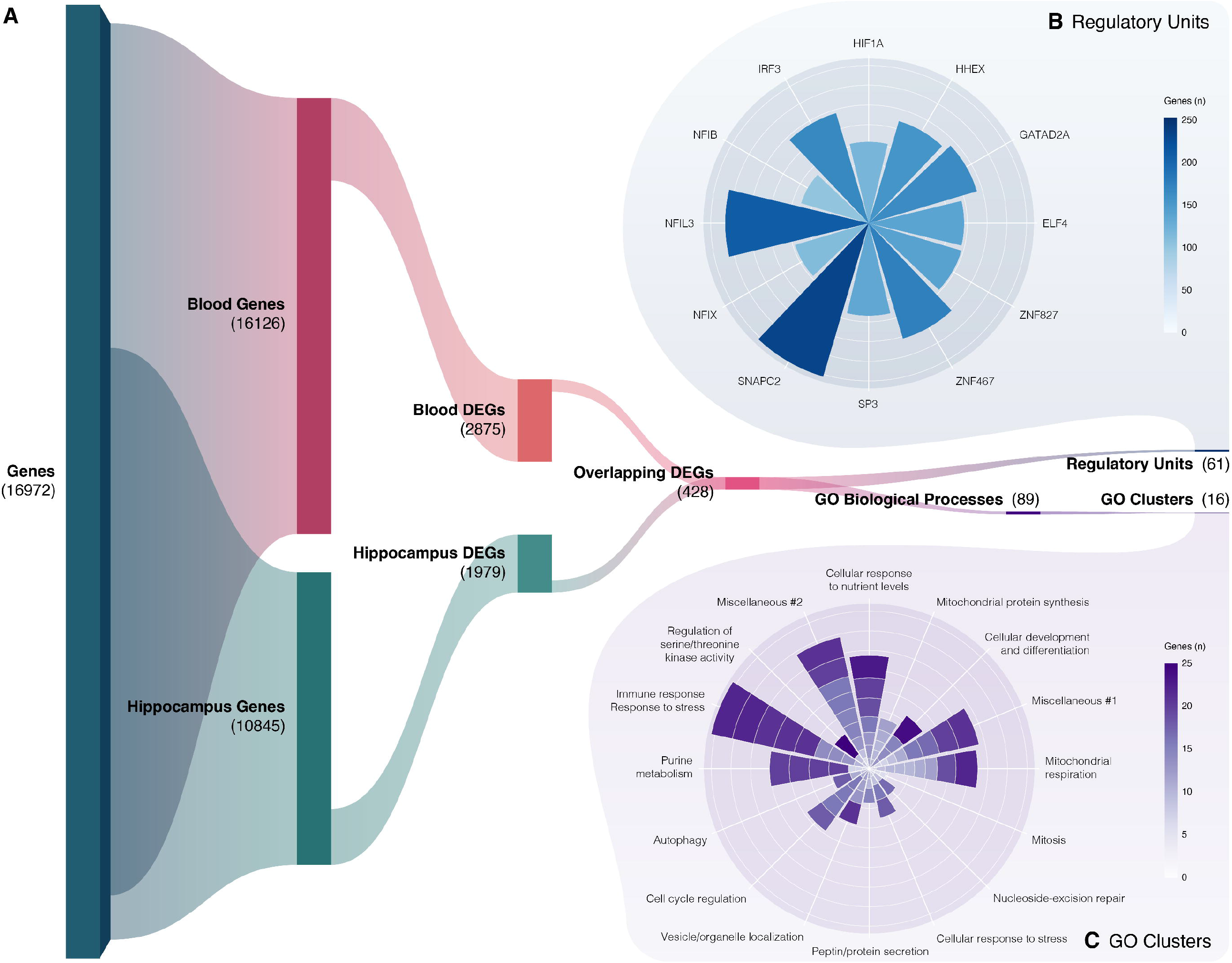
Sankey Flow Diagram representing the Dimension Reduction Strategy. (A) Blood and hippocampus transcriptomic data were obtained from the available cohorts. For each tissue, the Differential Expression Analysis (DEA) was performed comparing Cognitively Unimpaired (CU) and Cognitively Impaired (CI) individuals to generate Differentially Expressed Genes (DEGs). Considering the DEGs from blood and hippocampus, the ones that were consistently altered between tissues were selected (overlapping DEGs). The Gene Ontology (GO) Biological Processes were generated through the Functional Enrichment Analysis (FEA) of GOs using overlapping DEGs between blood and hippocampus. The semantic similarity method was used to group GO biological process terms into clusters of similar functional relevance (GO clusters). Regulatory Units of transcriptional factors (regulons) were generated using the same overlapping DEGs. (B) The Sunburst plot of Regulatory Units shows the number of genes for 12 regulons selected considering the tail-end quartiles (10% and 90%) of the enrichment score. (C) Given that the GO clusters are composed of multiple GOs, each segment in the Sunburst plot of GO clusters shows the number of genes for each GO that comprise the specific cluster.

Next, we used semantic similarity to group GO biological process terms into clusters of similar functional relevance. This dimension reduction strategy resulted in 16 clusters (**Figure 2**). **Figure 3A** shows these clusters in a network that represents node sizes proportional to cluster sizes (number of terms in the cluster) and edge width to the similarity score returned by the GOSemSim algorithm (see *Methods*; network average degree [sd] = 3 [2.07]). We observed that the clusters associated with *‘regulation of serine/threonine kinase activity’* and *‘DNA repair and metabolism’* had the highest connectivity metric (degree = 6) and degree/size ratio (degree/size = 2). Thus, albeit containing fewer terms than other clusters, these two clusters had the largest influence over the network. **Figure 3B** shows the heatmap of the semantic similarity matrix based on the GO terms and the resulting clusters of terms obtained from GOSemSim. Details about the clusters are reported in **Supplementary Table 6**. For the blood datasets, we also used reverse engineering of transcriptional networks to infer groups of regulatory units under the control of TFs. Similar to GO functional enrichment analysis and semantic similarity, the aim was to collapse groups of altered genes into biological modules. The MRA accomplishes this by identifying the regulatory units of TFs most enriched with DEGs. We observed 34, 92, and 41 regulatory units statistically altered in ADNI, GSE63063, and GSE97760 blood datasets, respectively (**Supplementary Table 7**). Sixty-one regulatory units were significantly enriched with DEGs in at least ⅔ of the datasets explored, and 12 were altered in all three. **Figure 4A** shows gene set enrichment analyses indicating the phenotypic associations (CI versus CU) of the top 10 regulons altered in the [^18^F]FDG-PET brain imaging. We also evaluated the GO functional enrichment of each top 10 regulons and mapped the results in **Figure 4B** as a network. This result showed that the genes inferred for most regulons are associated with biological processes. However, the regulatory units of GATA1 and ZNF358, for example, had mixed terms and were mainly related to cellular component terms. Interestingly, the ZNF653 regulatory unit is more connected with GOs related to energetic metabolism and protein kinase activity. Finally, we did not observe significant enrichment of GO terms for the ZZZ3 regulatory unit.

**Figure 3.**
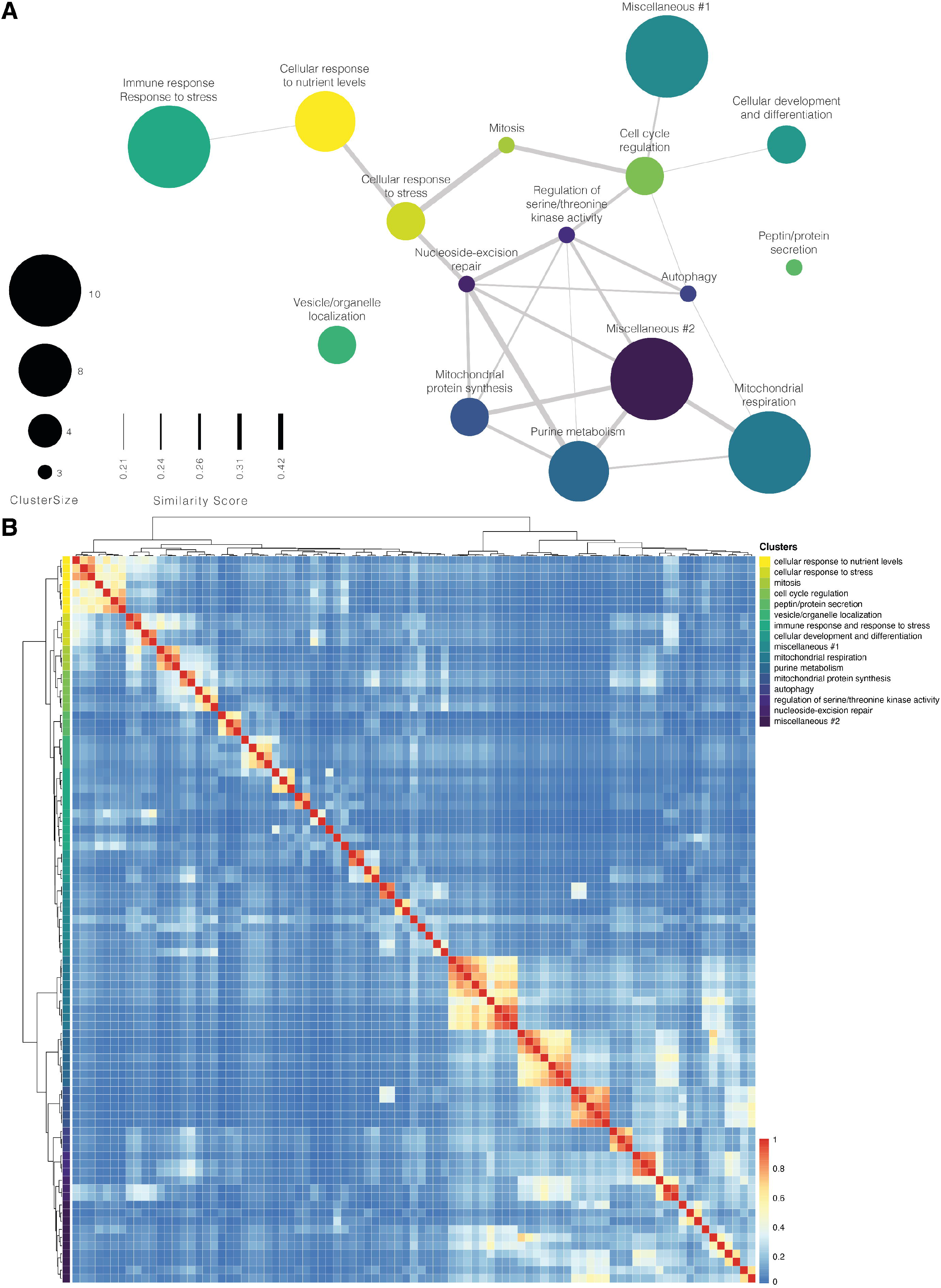
Functional Enrichment of Gene Ontology Biological Processes. (A) Gene Ontology (GO) semantic similarity network. GO biological process (GOBP) terms were clustered by semantic similarity and networked mapping cluster sizes to nodes and cluster similarity coefficients to edges. (B) Semantic similarity matrix heatmap of GOBP terms.

**Figure 4.**
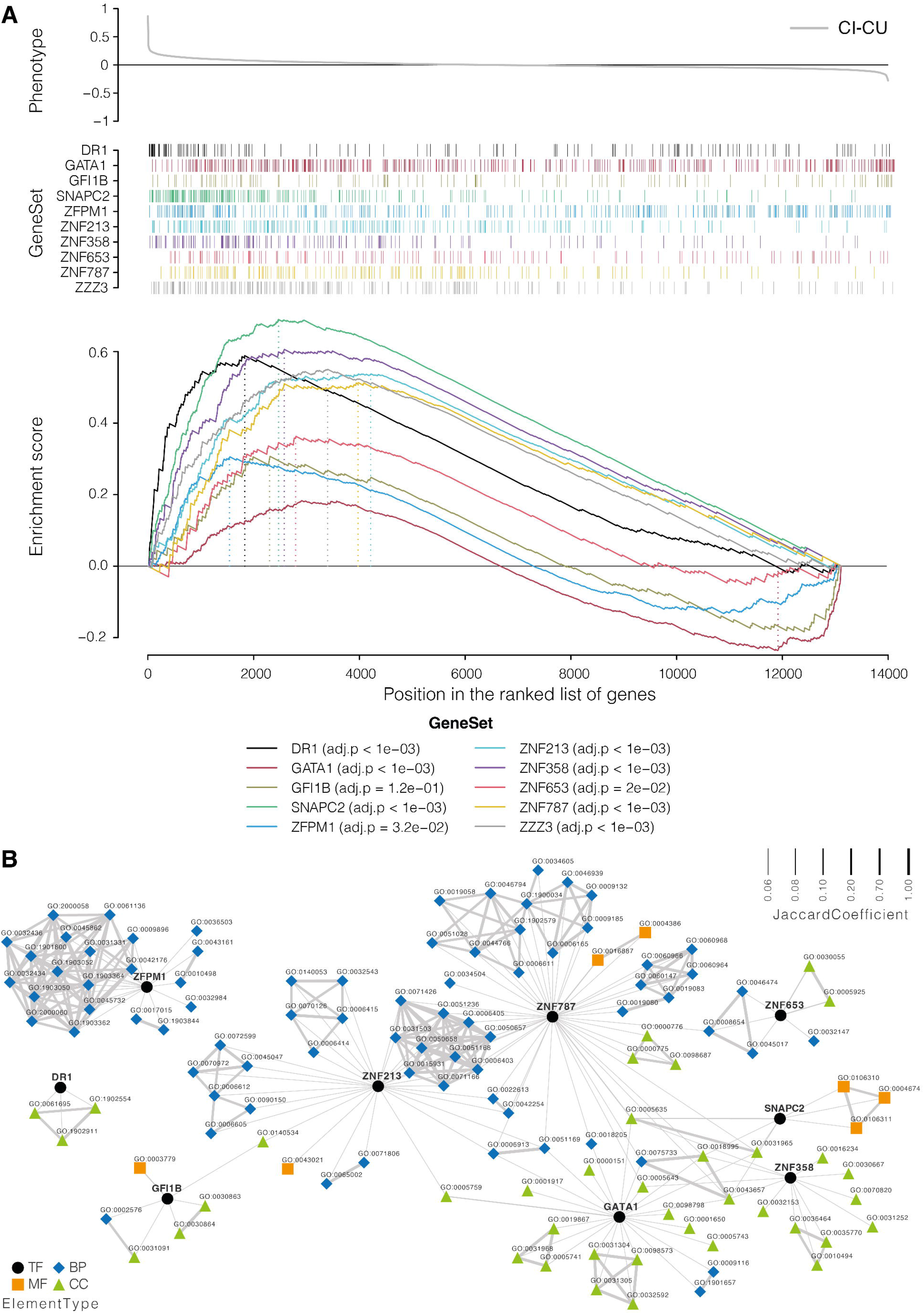
Enrichment of Regulatory Units of Transcription Factors. (A) Gene set enrichment analysis of top 10 regulatory units associated with brain imaging alterations. For each regulon, the gene set association was evaluated against Cognitively Impaired (CI) versus Cognitively Unimpaired (CU) phenotypes. (B) Gene Ontology (GO) functional enrichment of the top 10 regulons associated with brain modification. Each altered regulon was evaluated for GO enrichment and networked mapping of the nodes to different edges to the gene overlap between GO terms (Jaccard Coefficient). TF = transcription factors; MF = molecular function; CC = cellular component; BP = biological process.

With the 16 GO clusters obtained from semantic similarity analysis, we performed voxel-wise correlations testing the topographical associations between [^18^F]FDG-PET and each GO cluster enrichment score. Out of the 16 t-statistical maps obtained, the cluster involved in the regulation of protein serine/threonine kinase activity showed the most significant associations (t-value > 2.6, peak-t = 4.86, p-value = 0.01]) with cerebral glucose metabolism measured with [^18^F]FDG-PET (**Figure 5A**). Furthermore, the association between this cluster and the brain [^18^F]FDG-PET signal was more prominent in the precuneus’ gray matter (59.25% left, 68.63% right), medial frontal gyrus (52.76% left, 14.01% right), medial frontal-orbital gyrus (51.39% left, 11.58% right) and cingulate region (46.60% left, 18.46% right) (**Figure 5B**), regions highly vulnerable in AD.

**Figure 5.**
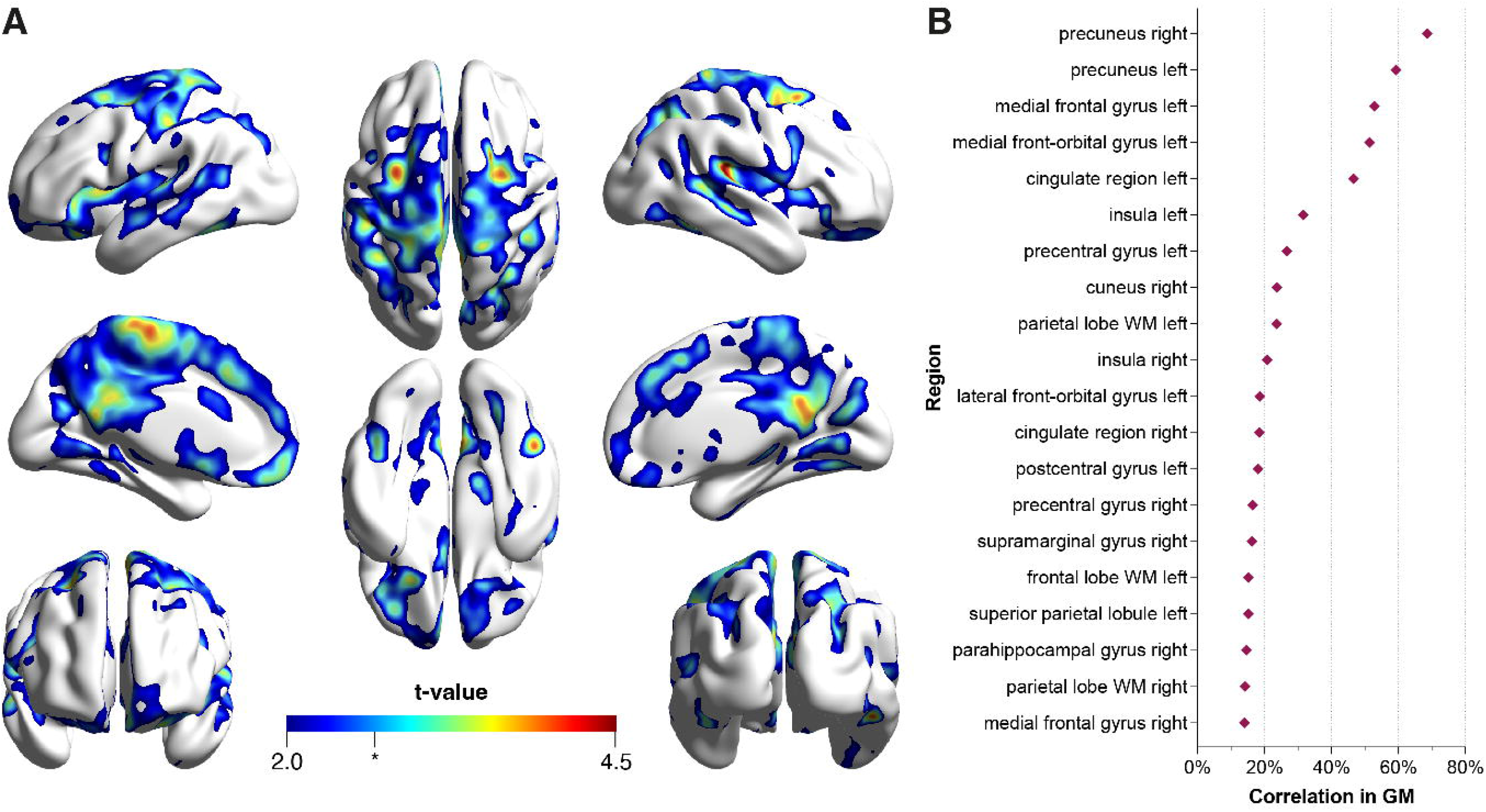
Voxel-wise correlation between the enrichment score of the Gene Ontology cluster related to the regulation of protein serine/threonine kinase activity and [^18^F]FDG-PET. (A) T-statistical map from the generalized linear regression model and (B) the top 20 brain regions with more statistically correlated voxels in the gray matter. t-value=2.0, DF=220, p-value=0.05; * tvalue=2.6, DF=220, p-value=0.01.

Similarly, we also performed voxel-wise linear regressions to investigate the spatial association between [^18^F]FDG-PET and the regulatory units enriched with DEGs. Among the 61 regulatory units, the regulatory unit of ZNF653 was the most closely associated with brain glucose metabolism (t-value > 2.6, peak-t = 3.90, p-value = 0.01]) (**Figure 6A**). Similarly, the association between this regulatory unit with [^18^F]FDG-PET signal was most observed in regions vulnerable to AD, such as the precuneus’ gray matter (24.24% left, 39.51% right), medial frontal gyrus (17.26% left), medial frontal-orbital gyrus (12.50% left) and precentral gyrus (9.08% left, 8.28% right) (**Figure 6B**).

**Figure 6.**
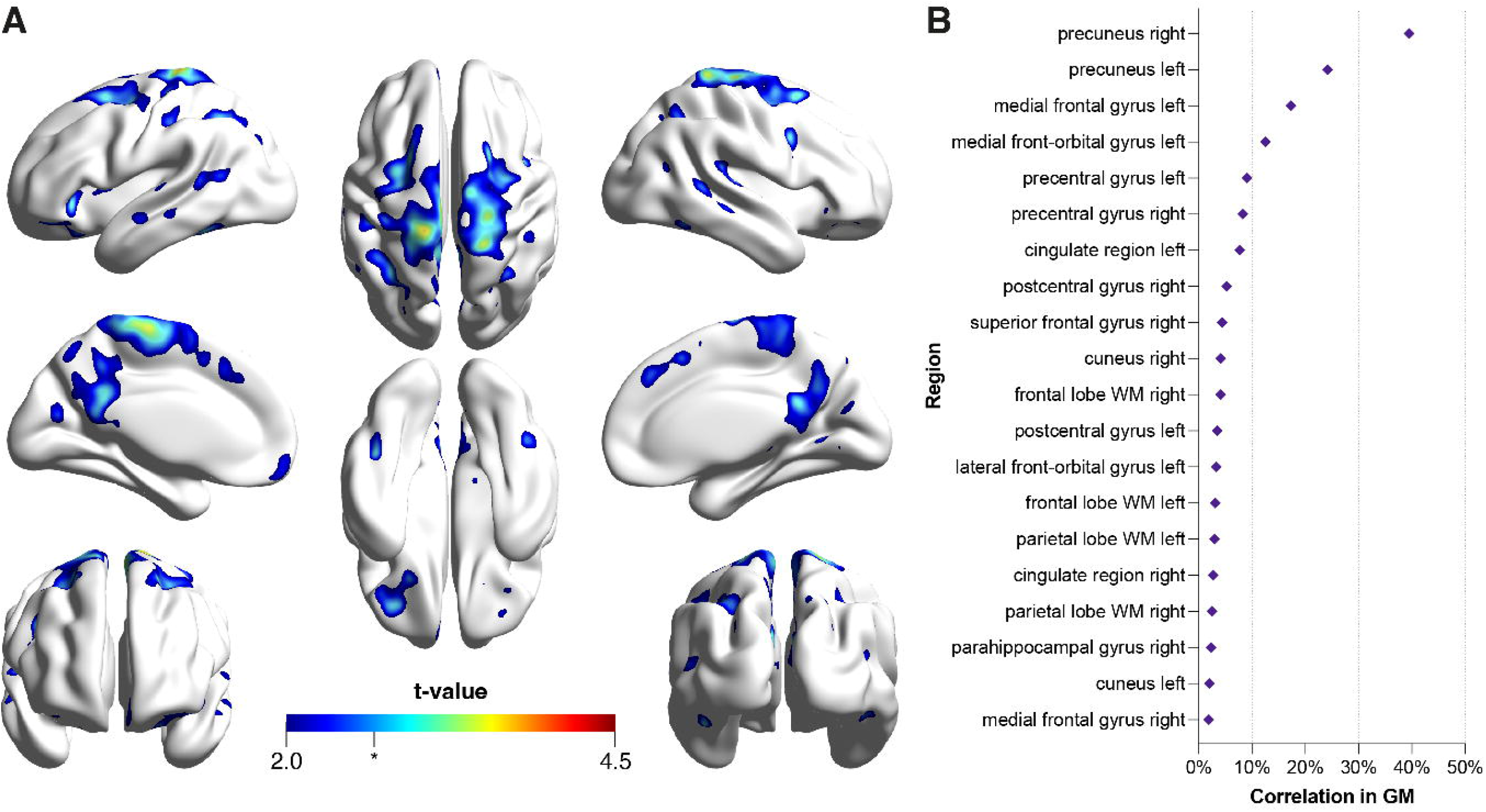
Voxel-wise correlation between the enrichment score of the regulatory unit ZNF653 and [^18^F]FDG-PET. (A) T-statistical map from the generalized linear regression model and (B) the top 20 brain regions with more statistically correlated voxels in the gray matter. t-value=2.0, DF=220, p-value=0.05; * tvalue=2.6, DF=220, p-value=0.01.

## DISCUSSION

In the present study, we developed a pipeline composed of two modules that integrates omics data with PET imaging. The first module involves omics analysis, composed of dimension reduction through the differential expression analysis, followed by functional enrichment analysis using the GO semantic similarity, or transcriptional network analysis, responsible for altered regulatory unit selection. The second module is imaging integration, where PET imaging is associated with statistically altered GO clusters and regulatory units. With this novel approach, we identified a GO cluster (regulation of protein serine/threonine kinase activity) and a regulatory unit (ZNF653) associated with changes in glucose metabolism in AD brain vulnerable regions.

The semantic similarity analysis of the GO biological processes revealed clusters of terms altered in AD that are significantly associated with [^18^F]FDG-PET imaging. Applying a reverse engineering strategy, we identified ten transcription factors and their regulatory units potentially acting as master regulators in our analysis and significantly associated with cerebral glucose metabolism. The [^18^F]FDG-PET is an essential modality for detecting functional changes in AD, clinically helpful in identifying changes in early AD and differentiating AD from other causes of dementia. The [^18^F]FDG-PET imaging has been used as a biomarker of neurodegeneration, with the source of signal claimed to be derived from neurons, astrocytes, and, more recently, microglia (42–44). This newly proposed integration method can help in the biological interpretation of [^18^F]FDG-PET, providing essential insights into understanding AD-associated neurodegeneration.

Our newly proposed integration method revealed that the GO cluster representing serine/threonine protein kinase activity was closely associated with [^18^F]FDG-PET metabolism. By adding phosphate groups quickly and dynamically, kinases can coordinate and control complex cellular functions, such as energy production, cell growth, and survival. Our brain is composed of hundreds of protein kinases, with a few already shown to lead to the spread of aberrant signaling in AD (45–47). In this sense, kinases such as Akt, extracellular signal-regulated kinase 1 and 2 (ERK1/2), cAMP-dependent protein kinase (PKA), glycogen synthase kinase-3β (GSK-3β), p70S6 kinase, and cyclin-dependent protein kinases 5 (Cdk5) were found with increased expression or activity in the AD brain. The hyperactivation of these kinases can lead to abnormal tau phosphorylation, amyloid production, apoptosis, and neuroinflammation. For instance, an increase in GSK-3β activity was observed in individuals with familial AD mutation of amyloid precursor protein (Swedish751) (46).

Our findings showed that brain glucose hyper, indexed [^18^F]FDG-PET imaging, correlates to TFs related to zinc-finger proteins. Specifically, among the top 10 regulatory units with the most association with [^18^F]FDG-PET, seven of them are from the zinc-finger family. The zinc-finger units regulate gene expression at the transcriptional and translational levels and are highly expressed in the brain (48). It has already been demonstrated that zinc-finger genes are associated with pathological changes in AD and regulate the expression of AD-related genes that are upstream in the production of hyperphosphorylated tau (49). This could explain the positive association when integrating [^18^F]FDG-PET and zinc-finger-related regulatory units enriched with DEGs.

We did not find any shared GO between the cluster representing serine/threonine protein kinase activity with the top 10 enriched regulatory units. However, the brain regions that showed significant correlations with zinc-finger genes are included in the regions that correlate with the serine/threonine protein kinase activity cluster. Even without sharing the same GO terms, this overlap could be interpreted as having more than one altered function related to glucose metabolism. Interestingly, the zinc-finger ZNF653 has 52 predicted phosphorylation sites, nine confirmed in humans, and five serine/threonine (50,51). This way, one could argue that the serine/threonine kinase may be modulating the activity of these TFs.

A biological definition of AD has recently been proposed solely based on AD biomarkers (52). Although it reflects fundamental advances in the development of neuroimaging and fluid biomarkers in AD research, a significant limitation of implementing this framework in clinical practices is the low predictive accuracy of this system (53). For instance, many Aβ and tau-positive CU individuals remain cognitively stable for their entire lifetime (53), suggesting that pathophysiological processes beyond Aβ and tau pathologies must be considered. Here, we detected significant biological pathways abnormalities in AD patients, which has the potential to provide insights into AD pathophysiology. Of note, the integration method proposed here is a novel tool that can be applied using multiple PET imaging tracers (*e*.*g*., Aβ PET, tau PET, TSPO PET), as well as adapted and used in the context of different neurological diseases and different omics modalities.

## CONCLUSION

We developed a strategy to integrate modular structures obtained from transcriptomic data, e.g., clusters of GO biological processes terms and regulatory units of TF, with [^18^F]FDG-PET imaging. Our results identified associations between protein serine/threonine kinase activity-related GO cluster and zinc-finger-related regulatory units with [^18^F]FDG-PET brain metabolism in AD.

## Data Availability

All data produced in the present study are available upon reasonable request to the authors.

## DECLARATIONS

### Ethics approval and consent to participate

The ADNI study was approved by the Institutional Review Boards of all participating institutions, and informed written consent was obtained from all participants at each site.

### Consent for publication

Not applicable.

### Data and code availability

All datasets used in this manuscript are publicly available in the referenced repositories. Further information will be readily available upon request.

### Author Contributions

GP: conceptualization, data curation, formal analysis, data interpretation, figures, writing – original draft, and writing – review & editing.

MAB: conceptualization, data curation, formal analysis, data interpretation, figures, writing – original draft, and writing – review & editing.

BB: data interpretation, figures, writing – original draft, and writing – review & editing.

PCLF: data interpretation, writing – original draft, and writing – review & editing.

JPFS: data interpretation, writing – original draft, and writing – review & editing.

FZL: writing - review & editing.

DOS: writing – review & editing.

PR-N: writing – review & editing.

BZ: supervision, writing – review & editing.

TAP: supervision, writing – review & editing.

ERZ: conceptualization, funding acquisition, supervision, writing – original draft, and writing – review & editing.

## Acknowledgments

Data collection and sharing for this project were funded by the Alzheimer’s Disease Neuroimaging Initiative (ADNI) (National Institutes of Health Grant U01 AG024904) and DOD ADNI (Department of Defense award number W81XWH-12-2-0012). ADNI is funded by the National Institute on Aging, the National Institute of Biomedical Imaging and Bioengineering, and through generous contributions from the following: AbbVie, Alzheimer’s Association; Alzheimer’s Drug Discovery Foundation; Araclon Biotech; BioClinica, Inc.; Biogen; Bristol-Myers Squibb Company; CereSpir, Inc.; Cogstate; Eisai Inc.; Elan Pharmaceuticals, Inc.; Eli Lilly and Company; EuroImmun; F. Hoffmann-La Roche Ltd and its affiliated company Genentech, Inc.; Fujirebio; GE Healthcare; IXICO Ltd.; Janssen Alzheimer Immunotherapy Research & Development, LLC.; Johnson & Johnson Pharmaceutical Research & Development LLC.; Lumosity; Lundbeck; Merck & Co., Inc.; Meso Scale Diagnostics, LLC.; NeuroRx Research; Neurotrack Technologies; Novartis Pharmaceuticals Corporation; Pfizer Inc.; Piramal Imaging; Servier; Takeda Pharmaceutical Company; and Transition Therapeutics. The Canadian Institutes of Health Research is providing funds to support ADNI clinical sites in Canada. Private sector contributions are facilitated by the Foundation for the National Institutes of Health (www.fnih.org). The grantee organization is the Northern California Institute for Research and Education, and the study is coordinated by the Alzheimer’s Therapeutic Research Institute at the University of Southern California. ADNI data are disseminated by the Laboratory for Neuro Imaging at the University of Southern California.

